# The true case fatality of COVID-19: An analytical solution

**DOI:** 10.1101/2020.05.17.20104554

**Authors:** Syamantak Khan

**Affiliations:** Department of Radiation Oncology, Stanford University

## Abstract

The exact risk of dying from COVID-19 has remained elusive and a topic of debate. In this study, the observed case fatality rates of 46 different countries are hypothesized to be dependent on their testing rates. An analytical test to this hypothesis suggests that the case fatality rate of COVID-19 could be consistent to a certain degree across all countries and states. The current global fatality rate is estimated to be around 1% and expected to converge between 1-3% when the pandemic ends. This model can be helpful to estimate the true infection rate for individual countries.

---

The case fatality rate (CFR) of COVID-19 is defined as the ratio of reported deaths from SARS-CoV-2 infection to the number of infected patients. The case fatality of COVID-19 and its variation across different countries remains debatable (*1-4*). In many countries, the case fatality even varies widely across different states (*5*). The observed inconsistency of CFR has been associated with age(*3*), co-morbidity(*6*), the efficiency of the healthcare system(*7*), three different viral strains(*8*), regional weather(*9*), and even vaccination schedule(*10*). However, none of these parameters are probably sufficient to explain the variation with mathematical precision.

To accurately calculate the fatality rate, it is necessary to correctly estimate the number of people infected by the virus, i.e., the denominator of the CFR ratio. (*2*). But, a salient feature of COVID-19 disease is its mild and asymptomatic nature in a large number of infected patients, who can carry and spread the infection to other healthy susceptible individuals. Data from recent studies suggest 25%-70% carriers of SARS-CoV-2 can be asymptomatic or show very limited symptoms (*11, 12*). Therefore, the efficiency of COVID-19 detection or testing is critical. Accurate detection not only helps isolate the infected patients, but it also allows accurate documentation of the infection and fatality rate. However, the testing infrastructure and policies also widely vary across nations, once again giving rise to an inconsistency in the fatality rate. This letter is intended to illustrate an intriguing relationship between the case fatality and testing rate which is gradually emerging from the COVID-19 dataset worldwide. A mathematical correlation function was derived using a simple model of infection transmission.

As only a very small percent of infected people develop severe symptoms, the large number of mild and asymptomatic patients can be missed easily. To overcome this challenge, the detection strategies (Covid-19 testing) follow an efficient algorithm. The probability of infection transmission from one particular individual gradually decreased from the close-contacts (persons who are close, or came close to the infected individual) to the far-contacts (persons who were physically distant from the infected individual). The probability of the spread of the infection approaches to zero for someone who is located at a large distance from the infected individual. Therefore the detection of COVID-19 infection is largely performed by a contact tracing method, mostly to maximize the efficiency of detection. In this method, when a patient (symptomatic or asymptomatic) is tested positive, all the individuals who came in close contact with that person in the last 10-14 days are tested with the highest priority. Thus, the detection algorithm is optimized by adapting the same pattern of the infection-transmission (see supplementary note Q1).

It is fundamentally important to accurately model the true detection efficiency in a population to know the true outbreak size and subsequently develop more efficient testing strategies. The chance of the contracting infection from one infected person can be mathematically modeled with a probability distribution function, which gradually decreases from the close-contacts to the far-contacts. As the spread of infection occurs continuously and independently at a constant average rate, the probability distribution is assumed to be exponentially decaying from close-contacts to far-contacts according to the following equation (Figure 1):

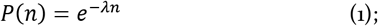

where *P(n*) is the probability of contracting the infection for *n^th^* contact person and *λ* is a constant representing the rate of decay. The physical distance of a contact person from the infected individual increases equivalently with the numerical value of *n*. The unit cell (or the building block) of this model includes only a single infected person at the center and the people who can contract the infection from that person directly. In reality, the same unit cell repeats itself for secondary, tertiary, and subsequent infections. Thus, like a growing crystal in a crystallization process, the infection keeps spreading within a population by repeating the unit-cell of infection following the same probability distribution function (equation 1).

**Figure 1:**
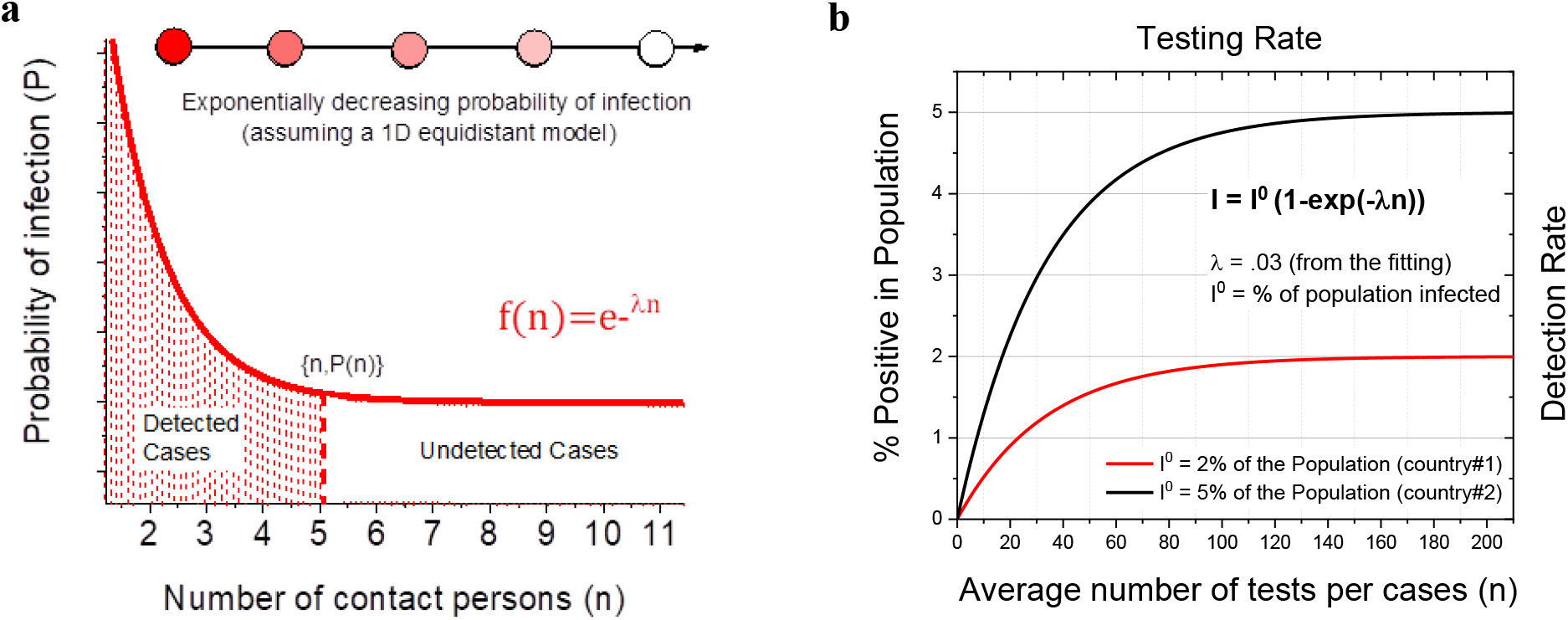
The mathematical model of COVID-19 detection. (a) A probability function of infection transmission from a single infected individual. The function can also indicate the probability of finding the n^th^ person COVID-19 positive while contact tracing. The area under the curve represents the total number of infected persons. The shaded area indicates the detected fraction of the population when only n number of contacts are tested for the disease. (b) Detection rate as a function of the testing rate in a population. The detected % positive rate initially increases with increasing testing rate but eventually saturates to the true % of infection within the population.

The total number of infection (*I*^0^) within a unit-cell can be estimated as (total area under the curve in figure 1a):

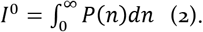

However, in an actual scenario, only a defined number of individuals can be detected by testing. Thus, the number of detected (*I*) infection within a unit-cell can be estimated as (shaded area under the curve in figure 1a):

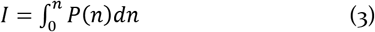

where *n* is the number of individuals tested per detected case through contact tracing, a number which widely varies across countries. The value *I* is equal to the area under the curve as illustrated in figure 1. The mathematical expression of *I* can be derived as a function of *n* by solving equations 1-3. Therefore, the detection efficiency (Φ) can be estimated as the ratio of the detected number to the actual number of infection within the unit cell:

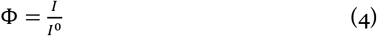

It is important to note that this ratio is dimensionless and independent of the total number of population in a region. In other words, this relationship is not limited to the unit-cell but applies to the whole population. Therefore the detection rate of COVID-19 within a population can be modeled using the relation given in equation 4. The simulated curve in figure 1b illustrates that at a given time in a region, the % detection of infected individuals increases when the testing rate is increased (see supplementary note Q2), but eventually saturates when most of the infected people are already detected within that population (Φ ≃ 1). The numerical value of exponential decay constant (*λ* =.03) was derived from fitting a dataset, which will be discussed later.

If *D* is the number of deaths from the infection within the unit-cell, then the actual CFR (δ^0^) and observed CFR (δ) can be defined as:

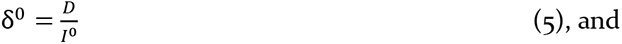

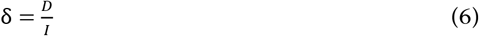

respectively.

Solving equation 1-6, the following relation can be established between the observed case fatality (dependent variable) and the number of testing performed per detected case (independent variable):

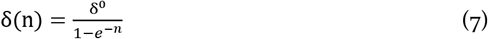

The uncertainty of the above function (∆*δ*) can be determined using the propagation of uncertainty from *I* as follows:

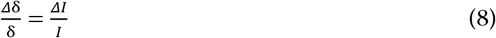

Where ∆*I* is the uncertainty of *I* and can be calculated from equations 1 and 3.

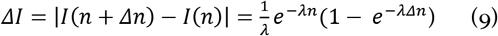

Solving equation 7-9, the uncertainty of observed CFR is found to be

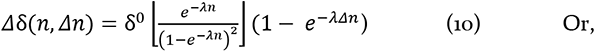

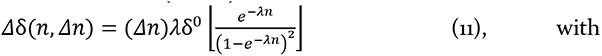

approximation (Taylor expansion).

Similar to detection efficiency, the equation 7-9 also holds for the whole population outside the unit-cell. To test these correlations within the existing datasets of COVID-19 pandemic, it is hypothesized that the actual or true fatality rate of COVID-19 is consistent (δ^0^ is a constant) worldwide. The values of δ(n) (total deaths/total cases) and n (total tests/total cases) were calculated for 46 different countries (Table S1, data collected from worldometers.info). The δ(n) vs *n* plot was fitted using equation 7 to determine the value of δ^0^. The fitting and the relative location of some countries are illustrated in Figure 2. The residual analysis is shown in the supplementary information (Figure S1). The goodness of fit can be further improved by excluding a few countries like Brazil and Italy who appear to be the outliers in the plot. Yet, the hypothesis can be tested keeping the uncertainties in mind.

**Figure 2.**
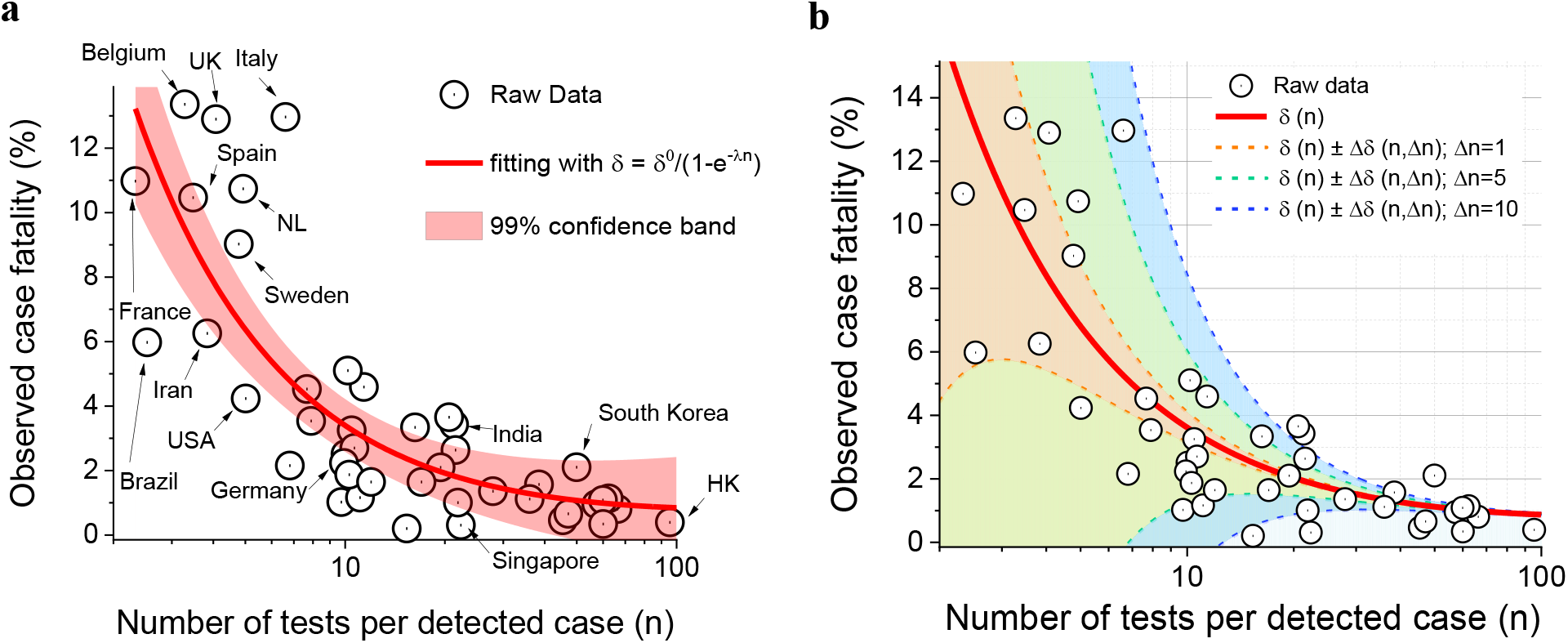
Correlation between the observed fatality rate and the average number of tests performed per detected case obtained from 46 countries. (a) The fitted line (red) represents the exponential relation stated in equation 7. The shaded area is the region of 99% confidence. Positions of several countries are indicated by arrows. (b) The regions of uncertainty (shaded area) of the observed case fatality (red line) exponentially reduce according to equation 11. The data points tend to converge to a fixed value of fatality rate with reduced uncertainty at a very high value of n.

According to equation 10, the uncertainty *∆δ* linearly increases with the value of ∆*n*, and decays exponentially as a function of *n*. The uncertainty regions are illustrated in Figure 2b for three different values of ∆*n*. The uncertainty analysis provides substantial supporting evidence to the hypothesis stated above. Conceptually, countries, where a large fraction of the screened people have been found to be contracted with the disease (SARS-CoV-2 positive), may have insufficient data to accurately calculate their fatality rate. The observed fatality rates in such countries could be overestimations and may fall within a broad range as well. In contrast, countries that have found most of the screened suspects to be negative for the disease (SARS-CoV-2 negative) have higher odds of converging into the accurate estimation of the fatality rate. The fitted value of δ^0^ suggest that the true case fatality of COVID-19 mathematically converges to 0.8% (δ^0^ = 0.8 ± 0.6) with relatively small uncertainty (Figure 2b). This numerical value is very similar to the fatality rate among the individuals in the Diamond Princess cruise ship (0.99%), a unique case study that might have provided us a reliable estimation of the CFR of COVID-19 (*2*).

It must be mentioned here that the presence of outliers in equation 7 is plausible with a low testing ratio (*n*), as the cause of death may depend on a large number of parameters as mentioned before. Hence, the fatality rate may not be exactly the same for each and every country. For instance, the observed fatality rate of Italy would be expected around 5% from equation 7, but it is >12% in practice due to several other factors. In contrast, the observed fatality is lower than expected in Brazil. Although these variations are expected from the uncertainty equations, the accurate values for individual countries cannot be estimated using this model.

An important question, which should arise in this context is whether the CFR reflects the true severity of the disease. As there is a time-lag between recovery and death, the CRF may keep changing throughout the epidemic in each country and saturate to a certain value at the end. Additional considerations might be needed to account for the time delay between disease onset and death (or recovery) from COVID-19 infection for the assessment of the actual clinical severity of the disease. For that, one can also look at the fatality rate in closed cases (CCFR), which takes the relative speed between death and recovery into considerations. At a given time point CCFR is generally found to be higher than CRF due to the long time delay between infection and recovery (figure 3a, supplementary information figure S2). A fitting of CCFR with a similar method results in a global fatality rate of around 3% (δ^0^ = 2.7 ± 0.3). The CCFR is expected to gradually decrease and converge to the same value of CFR at the end of the pandemic. Thus, at any time point, both of these time-sensitive indices may inaccurately represent the clinical severity. Yet, they provide a reliable window for the true fatality range which gradually diminishes with the duration of the pandemic (figure 3b). This range of uncertainty would gradually reduce with time as the pandemic progresses.

**Figure 3.**
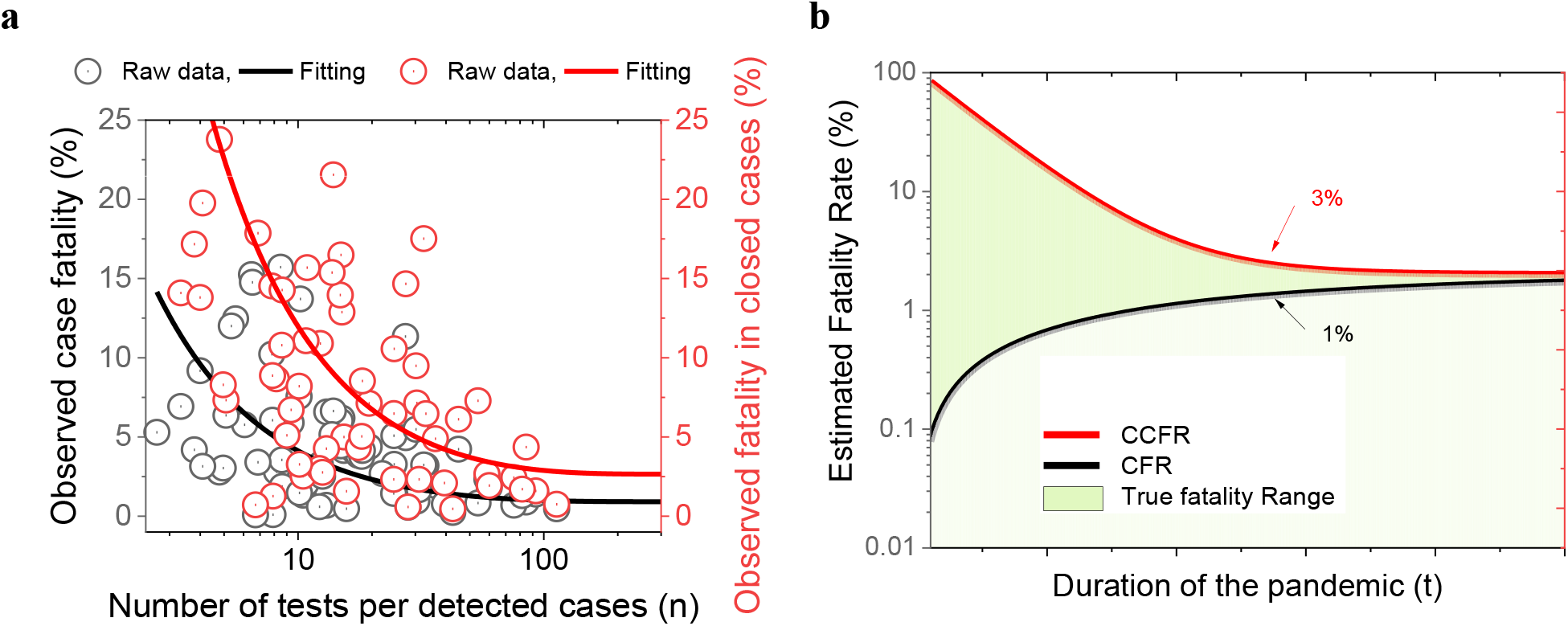
The two different types of fatality rate: Observed case fatality rate (CFR) and Observed closed-case fatality rate (CCFR) (a) The fitted lines show the difference between CFR (black) and CCFR (red). The CRF converges to 1% and CCFR converges to 3%. (b) Schematic diagram showing how CFR gradually decreases and CCFR gradually increases during the pandemic. They converge to the same value at the end of the pandemic. The uncertainty range for the true fatality rate is shown by the green shaded area.

One corollary which immediately manifests from this hypothesis is a possible route of estimation of the actual size of the outbreak. If it is assumed that most of the undetected patients recover from a mild or asymptomatic infection, then the current observed rate of recovery would also be an underestimation. According to the model discussed above, the outbreak size can be approximately estimated as a function of *n* using equation 1-4 (Figure 1b) and the fitted value of *λ* (Figure 4a). Assuming 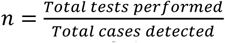, and λ = 0.03, equation 4 suggests that most of the countries may have already missed a significant portion of the infected population who might have recovered from a milder disease (Figure 4b). For instance, France, Brazil, Iran, and Belgium who have reported one of the highest rates (~30-50%, n≃2-3) of positive cases during Covid-19 detection, might have missed up to 75-90% of the true infected population till May 3, 2020. In contrast, Vietnam, UAE, Hong Kong, South Korea, or New Zealand who have reported the least rate of positive cases (~0.2-2%, n*≃*50-500) might have missed only 1-10% till May 3, 2020, according to this model. However, to estimate the number of recovered individuals from mild infection, it might need an in-depth formulation (Figure 4a) of the rate equations, which is outside the scope of this letter.

**Figure 4:**
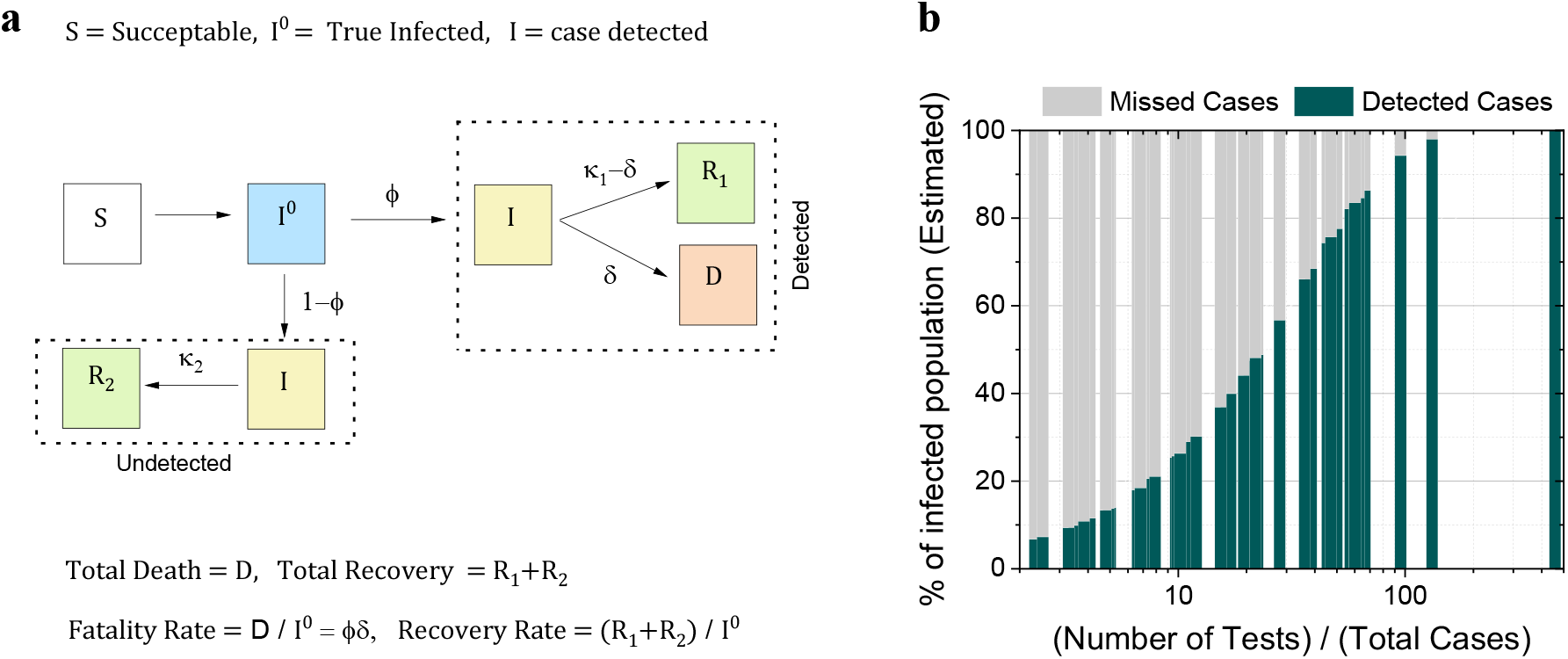
Detected and missed cases (a) Schematic diagram showing the true size of outbreak, recovery, and death. The proportion of undetected cases can be estimated as a function of detection efficiency (Φ). (b) The estimated percent of detected and missed cases as a function of testing rate across 46 countries. Most of the countries may have missed a significant proportion of cases according to the model.

So, in conclusion, the analytical approach presented in this letter shows that the widely variable observed fatality rate is a function of the testing rate or the average tests performed per case (see supplementary note Q3). However, the true case fatality rate has a fixed value between 1-3% in most of the countries. The methodology described here could be useful to estimate the true size of the infected population, which otherwise largely remains unknown. The accuracy of this simple mathematical approach can be improved further with additional parameters in future studies. This study attempts to reliably distill the data from various countries into a coherent mathematical formulation to estimate the global risk of mortality during the COVID-19 pandemic.

## Data Availability

Research data will be shared upon request to the corresponding author. The raw data were obtained from public databases.

https://www.worldometers.info/

https://www.covid19india.org/

## ASSOCIATED CONTENT

The supplementary information file contains table S1, Figure S1-2, and supplementary notes Q1-3.

## ACKNOWLEDGMENT

The author acknowledges Stanford University for the infrastructure and postdoctoral support. The author would like to thank his mentor Dr. Guillem Pratx, for providing valuable feedback on this paper. The author also thanks Dr. Soumya Poddar, Dr. Debarati Bandyopadhyay, and Dr. Taniya Sing for valuable discussions.

